# Optimising activity and diet compositions for dementia prevention: Protocol for the ACTIVate prospective longitudinal cohort study

**DOI:** 10.1101/2021.07.28.21261299

**Authors:** Ashleigh E. Smith, Alexandra T. Wade, Timothy S. Olds, Dorothea Dumuid, Michael J. Breakspear, Kate E. Laver, Mitchell R. Goldsworthy, Michael C. Ridding, Monica Fabiani, Jillian Dorrian, Montana McKewen, Bryan Paton, Mahmoud Abdolhoseini, Fayeem Aziz, Maddison L. Mellow, Clare E. Collins, Karen Murphy, Gabriele Gratton, Hannah A.D. Keage, Ross T. Smith, Frini Karayanidis

## Abstract

**Introduction:** Approximately 40% of late-life dementia may be prevented by addressing modifiable risk factors, including physical activity and diet. Yet, it is currently unknown how multiple lifestyle factors interact to influence cognition. The ACTIVate Study aims to 1) Explore associations between 24-hour time-use and diet compositions with changes in cognition and brain function; and 2) Identify durations of time-use behaviours and the dietary compositions to optimise cognition and brain function.

**Methods and analysis:** This three-year prospective longitudinal cohort study will recruit 448 adults aged 60-70 years across Adelaide and Newcastle, Australia. Time-use data will be collected through wrist-worn activity monitors and the Multimedia Activity Recall for Children and Adults (MARCA). Dietary intake will be assessed using the Australian Eating Survey food frequency questionnaire. The primary outcome will be cognitive function, assessed using the Addenbrooke’s Cognitive Examination-III (ACE-III). Secondary outcomes include structural and functional brain measures using Magnetic Resonance Imaging (MRI), cerebral arterial pulse measured with Diffuse Optical Tomography (Pulse-DOT), neuroplasticity using simultaneous Transcranial Magnetic Stimulation (TMS) and Electroencephalography (EEG), and electrophysiological markers of cognitive control using event-related potential (ERP) and time-frequency analyses. Compositional data analysis, testing for interactions between time-point and compositions, will assess longitudinal associations between dependent (cognition, brain function) and independent (time-use and diet compositions) variables.

**Conclusions:** The ACTIVate Study will be the first to examine associations between time-use and diet compositions, cognition and brain function. Our findings will inform new avenues for multidomain interventions that may more effectively account for the co-dependence between activity and diet behaviours for dementia prevention.

**Ethics and dissemination:** Ethics approval has been obtained from University of South Australia’s Human Research Ethics committee (202639). Findings will be disseminated through peer reviewed manuscripts, conference presentations, targeted media releases and community engagement events.

**Registration Details:** Australia New Zealand Clinical Trials Registry (ACTRN12619001659190).

**Strengths and limitations:**

- The ACTIVate Study will collect comprehensive measures of lifestyle behaviours and dementia risk over time in 448 older adults aged 60-70 years.
- Using newly developed Compositional Data Analysis (CoDA) techniques we will examine the associations between time-use and diet compositions, cognition and brain function.
- Data will inform the development of a digital tool to help older adults obtain personalised information about how to reduce their risk of cognitive decline based on changes to time use and diet.
- Recruitment will be focussed on older adults to maximise the potential of making an impact on dementia prevention in the next 10 years.
- Findings may not be generalisable to younger adults.

## INTRODUCTION

Dementia is one of the leading causes of death and disability in older adults [1, 2]. As the population ages, rates of dementia are expected to increase substantially. Worldwide, it was estimated that 46.8 million people were living with dementia in 2015 [3]. By 2050, this number is predicted to triple to 131.5 million [3]. In the absence of effective pharmacological treatments, and with up to 40% of late life dementia attributed to modifiable risk factors [4], prevention has moved to the forefront of dementia research.

### Activity and diet compositions

Lifestyle factors such as physical activity, sedentary time, sleep and dietary intake are known modifiable risk factors for dementia [5-8]. These factors have typically been researched as independent predictors of dementia risk and are either targeted separately in intervention studies or clustered together in multi-domain trials with little consideration of how they interact [9]. However, lifestyle factors are not independent of each other. In fact, physical activity, sedentary time and sleep are, by nature, co-dependent and constitute mutually exclusive and exhaustive parts of a 24-hour day. If time spent in one of these behaviours increases, time in other(s) must decrease to compensate. Time spent in these behaviours is also likely to be linked with dietary intake (for example, more time spent in front of television is associated with more snacking). In fact, dietary intake itself can also be conceptualised as a composition of distinct food groups which are co-dependent on each other [10]. That is, increased consumption of one food or food group may lead to relative compensatory changes across other foods or food groups. Thus, time-use behaviours and dietary intake should be considered as *interrelated compositional* predictors of dementia risk. To more fully understand their role in dementia prevention and to better design interventions, it is necessary to both characterise and account for their co-dependency.

Although it makes sense to consider all daily time-use behaviours and all dietary components together, epidemiologists have largely been unable to include them in the same analytical model due to the perfect multi-collinearity between parts of such compositions. Recently, an analytical approach specifically designed for the analysis of compositions, named compositional data analysis (CoDA) [11], has been applied to overcome the multi-collinearity issue of time-use [5, 6] and diet [12] composition data, and to enable the inclusion of all components in the same statistical model. CoDA expresses time spent in different activities or proportions of food groups as isometric log ratios, which can then be used as variables in standard statistical analyses. CoDA can also consider *sub-compositions*, subsets of the activity or diet composition (for example, waking hours, or vegetable intake). In these models, an extra term may be included in the regression representing the sub-compositional total (e.g., the total number of waking hours, or the total number of serves from vegetables). These analyses will tell us whether the mix of activities or dietary components in the sub-composition, independent of the total, is associated with some outcome.

A recent application of CoDA, called compositional isotemporal substitution, [13] models the health effects of re-allocating time from one activity domain to another (for example, swapping 30 min of sleep for 30 min of screen time). Compositional isotemporal substitution effectively generates dose-response relationships between activity compositions and health outcomes.

Likewise, it is possible to model the effects of changing the proportion and composition of food groups in the diet. Recent studies have applied CoDA and related methods to identify how proportions of macronutrients, micronutrients and food groups are associated with waist circumference, plasma lipids and lipoproteins, and glucose concentrations [10, 14]. Just as isotemporal substitution models the effects of time reallocations, isocaloric substitution can be used to model the effects of dietary reallocations while accounting for variance in total caloric intake.

### Mechanisms of cognitive change

It is well accepted that structural and functional brain changes begin manifesting asymptomatically years before dementia is diagnosed [15-17]. These changes may include accelerated brain atrophy [15], accumulation of protein aggregate in the brain [18], disrupted integrity of the cerebrovascular system [19], and failure of the brain to undergo plastic re-organisation, otherwise referred to as neuroplasticity [20]. Imaging techniques that characterise brain structure and function (Magnetic Resonance Imaging, MRI), map brain temporal dynamics (Electroencephalography, EEG), and measure cerebral arterial pulse (Diffuse Optical Tomography, Pulse-DOT) [21, 22], as well as non-invasive brain stimulation techniques (repetitive Transcranial Magnetic Stimulation, TMS) are now at the forefront of identifying and tracking pre-clinical brain changes in dementia research [9, 23, 24].

### Aims

The ACTIVate Study will explore how daily time-use and diet compositions are associated with cognition to inform dementia prevention. Iterative analyses of different activity and diet compositions will be used to identify “best days” — optimal combinations of activities and food groups associated with the greatest possible reduction in dementia risk. Together, these analyses will inform the co-design of an interactive tool that will estimate the predicted change in cognition for changes in time-use and/or dietary behaviour.

The ACTIVate Study will also include a secondary mechanistic focus, tracking brain structure and function longitudinally. This novel approach will enable us to model the benefits of modifying either time use or diet, or both concurrently. By tracking secondary imaging and neurophysiological measures longitudinally alongside traditional cognitive assessments, we expect to be able to detect subtle brain changes that may occur before behavioural differences become apparent.

Using longitudinal data collected from older adults over a period of three years, the ACTIVate Study will aim to:

1. Explore how 24-hour time use and diet compositions are associated with changes in cognition and brain function;
2. Identify the best durations of time-use behaviours and the best proportions and compositions of food groups to optimise cognition and brain function.

## METHODS

### Ethics

The ACTIVate Study was registered with the Australia New Zealand Clinical Trials Registry (ACTRN12619001659190) on November 27, 2019. Ethics approval was obtained from the University of South Australia and University of Newcastle Human Research Ethics Committees (202639). All procedures will be conducted in accordance with the Declaration of Helsinki.

### Study design

The ACTIVate Study will employ a 3-year prospective longitudinal study design, with objective assessments at baseline, 18 and 36 months.

### Participants and recruitment

Four hundred and forty-eight community-dwelling adults will be recruited from Adelaide, South Australia (n=224) and Newcastle, New South Wales (n=224). Participants will be aged 60 to 70 years and fluent in the English language. Exclusion criteria will include: current clinical diagnosis of dementia; Montreal Cognitive Assessment-Blind (MoCA-B) score <13/22; blindness, colour blindness or vision difficulties not corrected by glasses or contact lenses; major neurological or psychiatric diagnosis; known intellectual disability; major physical disability; previous head trauma resulting in a loss of consciousness of more than five minutes; current or previous alcohol or substance abuse or dependence; recreational drug use in the last three months; previous diagnosis of stroke or transient ischemic attack; cancer treatment affecting cognitive performance (e.g., chemotherapy) in the last five years. The following additional exclusion criteria will apply to MRI procedures: the presence of a pacemaker, metal implants, cochlear or other ear or body implants; the presence of metal fragments or other foreign bodies in the eyes, skin or body; a history of claustrophobia. Additional exclusion criteria will apply to participants in Adelaide undergoing TMS: current medications which may lower seizure threshold; a history of epilepsy, convulsions or seizures; a history of surgical procedures to the spinal cord or any spinal or ventricular derivations [25].

Using a rolling convenience sampling strategy, all participants will be enrolled over an 18-month period. Paper, radio and electronic advertisements will be disseminated across the Adelaide greater metropolitan area and the Hunter Region. Existing volunteer databases will also be utilised. Volunteers will first undergo a phone screening interview, during which the MoCA-B will be administered to screen against dementia. Screening for other exclusion criteria will also be completed. Volunteers who meet all eligibility criteria will be invited to participate and scheduled for Baseline assessments. To ensure similar inclusion/exclusion criteria are adhered to at both study sites a weekly eligibility meeting will be conducted to discuss any potential cases with an eligibility query.

### Measures

#### Primary Dependent Variable: Cognitive function

The Addenbrooke’s Cognitive Examination-III (ACE-III) is the primary outcome of the ACTIVate Study. The ACE-III is a brief cognitive screening tool that demonstrates high internal reliability (Cronbachs α = 0.88), as well as high sensitivity (1.00) and specificity (0.96) for the detection of dementia using the cut-off score of 88/100 [26]. For the purpose of the ACTIVate Study, the ACE-III score will be assessed as a continuous variable. The ACE-III assesses function across five cognitive domains: memory, attention/orientation, fluency, language and visuospatial. Our pilot data also indicate that the ACE-III is sensitive to intra-individual differences in daily activity patterns. To control for possible practice effects, all three Australian versions of the ACE-III will be used, with Version A administered at Baseline, Version B at 18 months and Version C at 36 months.

#### Primary Independent Variables

##### Daily time-use composition

Time-use data will be captured objectively using 24-hour 7-day accelerometry with Axivity AX3 wrist-worn triaxial devices. Triaxial accelerometers measure acceleration along three planes (vertical, anteroposterior and mediolateral) and have high predictive validity for physical activity intensity [27]. The AX3 has demonstrated a high level of accuracy in detecting and classifying time spent in physical activity [5] and has been validated for use in older adults [28]. Wrist-worn AX3 monitors have been chosen over lower limb-mounted devices due to evidence of higher levels of wear time compliance across a 24-hour period [29].

While wearing the AX3, participants will be asked to maintain a self-report record of sleep and wake times and to detail any periods of removal. Raw accelerations will be condensed into 60-s data epochs and categorised into either time spent in sedentary behaviour, light physical activity, moderate physical activity or vigorous physical activity, using pre-defined cut-points (46/93/418 for light, moderate and Vigorous physical activity, respectively) adjusted for 100 Hz sampling frequency [30, 31]. Sleep duration and disturbances will be classified using an algorithm developed by van Hees et al. [32], in conjunction with self-report record of non-wear, sleep and wake times.

Self-reported time-use data will be collected using the Multimedia Activity Recall for Children and Adults (MARCA). The MARCA is a computerised time-use recall tool containing over 500 activities linked to an energy expenditure compendium [33]. When completing the MARCA, participants will be asked to recall every activity they engaged in over the previous two days in 5-minute increments. Activity categories include ‘Passive Transport’, ‘Sports and recreation’, ‘Work’, ‘Self-Care’, ‘Household Chores’ and ‘Screen Time’. The MARCA has strong validity when compared to accelerometry data in adults (rho=0.72), and excellent test-retest reliability (ICC: 0.99 – 1.00) [34].

##### Diet Compositions

Diet composition will be operationalised as serves per day, between and within five food groups: 1) Fruits and vegetables; 2) Protein; 3) Fats 4) Cereals; 5) Discretionary foods. In the first step, we will determine the optimal proportion of each food group as part of the whole diet. The second step will identify the optimal composition of each food group. The compositions of each food group are presented in Table 1, and are informed by the Australian Dietary Guidelines [35] and previously published associations between dietary intake, cognitive function and dementia risk [36-39].

**Table 1.**
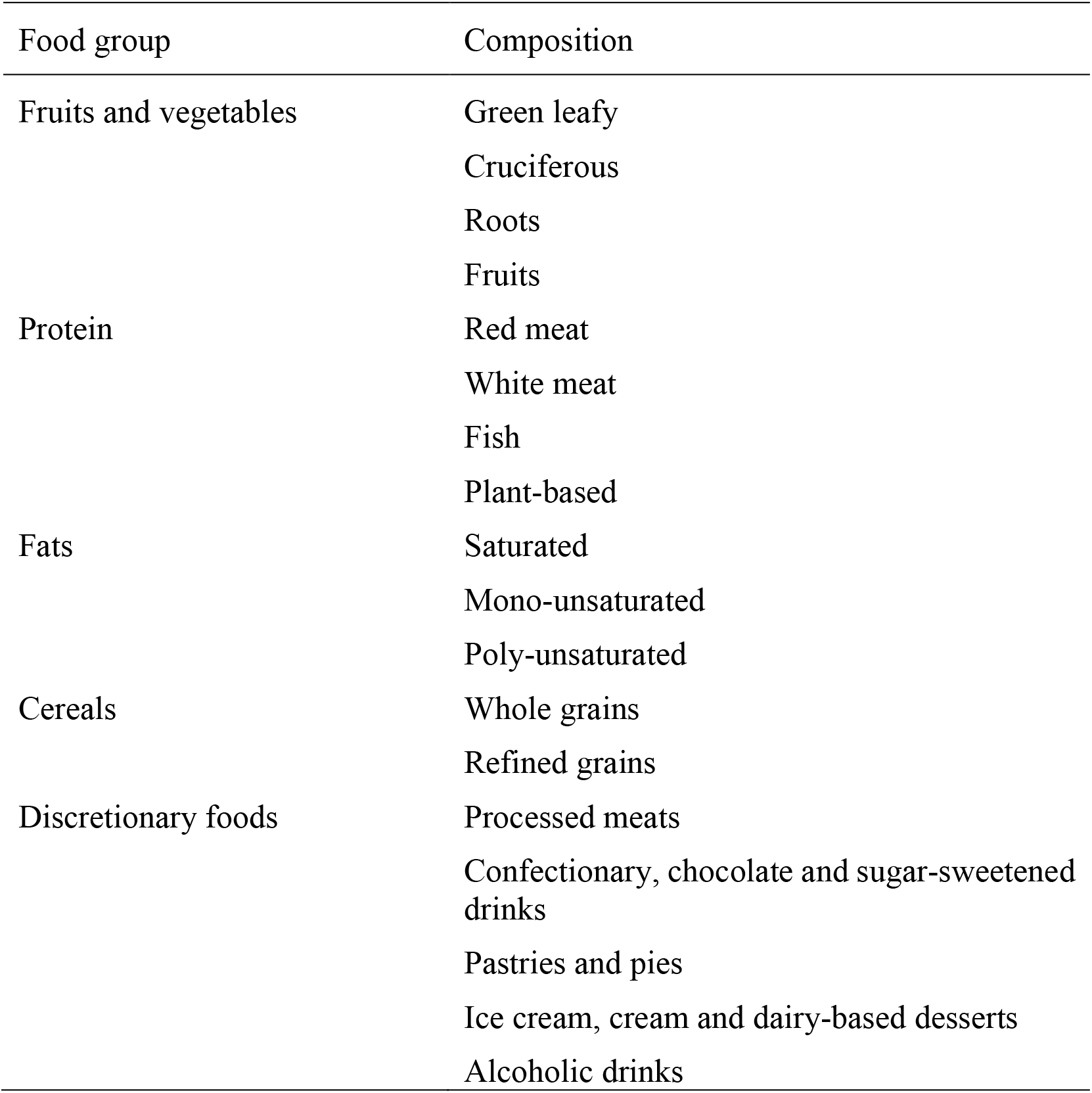
Food group compositions

Dietary intake will be measured using the Australian Eating Survey, a 120-item semi-quantitative food frequency questionnaire designed to capture the nutrient intake of Australian adults against a contemporary list of foods that represent the current Australian food supply [40]. Participants will be asked to report how frequently they consume different foods and food types over the past 6 months, with responses ranging from ‘Never’ to 4 or more times per day’, with some beverages up to ‘7 or more glasses per day’. Foods are organised into the following categories: drinks, breads and cereals, dairy food, main meals, sweets and snacks, fruit and vegetables. The Australian Eating Survey provides accurate estimates of nutrient intake for macro- and micronutrients which have been validated against 4-day weighed food records [40], as well as plasma carotenoids for fruit and vegetable intake [41] and erythrocyte fatty acids [42].

Consumption of oils, such as extra virgin olive oil, have been associated with reduced risk of cardiovascular disease and cognitive decline, and are therefore of interest to the current study [21-23]. To estimate oil intake, an additional dietary questionnaire will be administered. The structure of the dietary oils questionnaire is based on the polyunsaturated fatty acids questionnaire developed by Swierk et al [43]. Participants will be asked whether they consume different dietary oils and fats, including butter, butter blends, margarine, margarine blends, lard, vegetable oil, canola oil, sunflower oil, coconut oil and olive oil. If participants respond ‘Yes’, they are prompted to indicate how much of each oil they usually consume each day, ranging from ‘Less than ½ a tablespoon’ to ‘8 or more tablespoons’.

#### Exploratory Secondary Biological Mechanisms

##### Magnetic Resonance Imaging (MRI): Adelaide and Newcastle

The MRI protocol will include T1-weighted structural scan, T2 FLAIR, diffusion weighted imaging (dMRI) and resting-state functional MRI (rs-fMRI) and associated field maps (Table 2). T1-weighted images will be used for co-registration of dMRI, rs-fMRI and optical imaging data, individual targeting of the prefrontal cortex with TMS, as well as morphometric analysis of white and grey matter volume and parcellation of different brain area volumes. T2 FLAIR will enable the analysis of deep white matter hyperintensities, combining with the T1 weighted images to create artificial myelin burden contrasts. dMRI will be used to derive white matter tract structural connectivity and rs-fMRI for analysis of functional connectivity. MRI scans will be collected on a Siemens Skyra 3T scanner in Adelaide and a Siemens Prisma 3T scanner in Newcastle, both using a 64-channel head and neck coil. MRI protocols have been calibrated across sites to ensure comparable data quality given the differences in the gradient systems and capabilities of the two different scanners.

**Table 2.** MRI scan parameters, duration and resolution

| **Sequence name** | **Acquisition time (min:sec)** | **Matrix size (mm<sup>3</sup>)** | **Voxel size (mm<sup>3</sup>)** | **TE (msec)** | **TR (msec)** | **FA (degree)** | **TI (msec)** |
| --- | --- | --- | --- | --- | --- | --- | --- |
| **Site/Scanner:** HMRI (Newcastle), 3T Siemens PRISMA, 64 channel head and neck coil |  |  |  |  |  |  |  |
| **T1 MPRAGE** | 5:12 | 256×256×208 | 1×1×1 | 2.91 | 2300 | 9 | 900 |
| **T2 FLAIR** | 4:12 | 256×256×176 | 1×1×1 | 386 | 5000 | T2 Variable | 1800 |
| **Diffusion** | 8:52 | 122×122×74 | 2×2×2 | 84 | 4700 | 90/180 | - |
| **Rs-fMRI** | 8:05 | 86×86×60 | 2.4×2.4×2.4 | 33 | 829 | 53 | - |
| **Site/Scanner:** SAHMRI (Adelaide), 3T Siemens SKYRA, 64 channel head and neck coil |  |  |  |  |  |  |  |
| **T1 MPRAGE** | 5:12 | 256×256×208 | 1×1×1 | 2.90 | 2300 | 9 | 900 |
| **T2 FLAIR** | 4:12 | 256×256×176 | 1×1×1 | 386 | 5000 | T2 Variable | 1800 |
| **Diffusion** | 11:00 | 122×122×74 | 2×2×2 | 114 | 5700 | 90/180 | - |
| **Rs-fMRI** | 9:31 | 70×70×60 | 3×3×3 | 39 | 1360 | 53 | - |

##### Transcranial Magnetic Stimulation: Adelaide site only

Concurrent TMS and EEG (TMS-EEG) and repetitive TMS will be used at the Adelaide site to investigate the secondary mechanism of prefrontal neuroplasticity [44], at Baseline and 3-years. Participants will complete two sessions (at least 7 days apart), each consisting of a ∼2-hour TMS-EEG protocol. EEG will be recorded at rest and while stimulating the cortex to assess neuroplasticity. Single-pulse TMS will be applied using a hand-held figure-of-eight magnetic coil connected to a Magstim 200 stimulator (Magstim, Whitland, UK). A neuro-navigation system (ANTneuro visor2) loaded with individual structural MRIs will be used to localise the left superior frontal gyrus (SFG). Additionally, the neuro-navigation software will be used to calculate coil-to-cortex distance of the motor cortex and SFG. Continuous theta-burst stimulation (cTBS) will be delivered at 70% resting motor threshold, adjusted for the difference between coil-to-cortex distance between the motor hotspot and SFG [45]. To experimentally-induce neuroplasticity, an inhibitory repetitive TMS paradigm, cTBS (real or sham, with session order randomised), will be applied for 40 seconds to the left SFG. Changes in SFG excitability from pre to post cTBS will be quantified as changes in the amplitude of TMS-evoked potentials (TEPs) in the EEG signal. TEPs will also be recorded for an equivalent amount of single-pulse TMS stimulations to the left shoulder before and after cTBS, in order to measure peripherally-evoked potentials and minimise the contribution of sensory artifacts to TEPs [46]. Participants will be instructed to sit quietly, keep their eyes open, and look straight ahead during each TMS-EEG recording period, and a masking noise will be applied through earphones (< 70 dB in each ear) to minimise contamination of the EEG by the audible click of the TMS discharge.

##### Diffusion Optical Tomographic (DOT) imaging: Newcastle site only

Cerebral arteries and arterioles become less able to flexibly contract and dilate with changing metabolic demands and challenges (i.e., *cerebrovascular reactivity*) and their walls become stiffer (i.e., *arteriosclerosis or reduced arterial elasticity)*. This is particularly evident in the prefrontal cortex (PFC) [22], which supports cognitive control processes. Reduced reactivity and elasticity of the brain’s arteries are associated with structural changes in the corresponding cortex [23] and decline in associated cognitive abilities [47]. Pulse-DOT (cerebral arterial pulse measured with diffuse optical tomography) provides a direct measure of the regional reactivity and elasticity of cerebral arteries with *high spatial resolution* (by co-registration to individual anatomical MRI scan).

Cerebral arterial elasticity and arterial reactivity will be measured at the Newcastle site using non-invasive pulse-DOT and fNIRS measures recorded with an ISS Imagent device. Three montages (with four detectors and 32 sources each) will be used to cover the participant’s prefrontal cortex using a custom-made foam cap with 2 min recording per montage. The location of possible fibre positions across the scalp and fiducial points will be digitized (Polhemus 3D) to allow detector/source locations to be co-registered to the participant’s T1 anatomical MRI image. EKG (500 Hz, 1-100 Hz bandpass filter) will be used to identify the R wave and synchronise recording montages and locations. Pre-processing of the AC-intensity data will use measures established previously [27, 28]. We will record at rest to derive pulse relaxation function (PReFx), a regional measure of arterial elasticity which is sensitive to the status of cortical arteries whose downstream branches perfuse periventricular areas, where white matter hyperintensities are most common. We will also measure cerebrovascular reactivity by recording during a Breath Holding Task (BHT). which induces hypercapnia in the brain [48, 49] Changes in optical pulse amplitude will be obtained during difference phases of the BHT, to assess reactivity to the hypercapnia challenge, using parameters and trial structure previously established [50].

#### Secondary measures

##### Secondary cognitive outcomes

###### Neuropsychological assessments

The Cambridge Neuropsychological Test Automated Battery (CANTAB) will be administered to assess executive function, memory and processing speed domains that are known to be sensitive to age-related cognitive decline and dementia [7, 51]. Previous studies have demonstrated that these cognitive domains can be influenced by lifestyle factors such as physical activity and diet [52-55]. The chosen CANTAB tests include Motor Orientation Task (MOT); Paired Associates Learning (PAL); Verbal Recognition Memory (VRM), Immediate and Delayed; One Touch Stockings of Cambridge (OTS); Reaction Time (RTI); and Multitasking Test (MTT). Subtests of the NIH Toolbox (Oral Reading Recognition, ORR, and Picture Vocabulary (PV), will be administered to assess language. A summary of cognitive assessments is presented in Table 3.

**Table 3.**
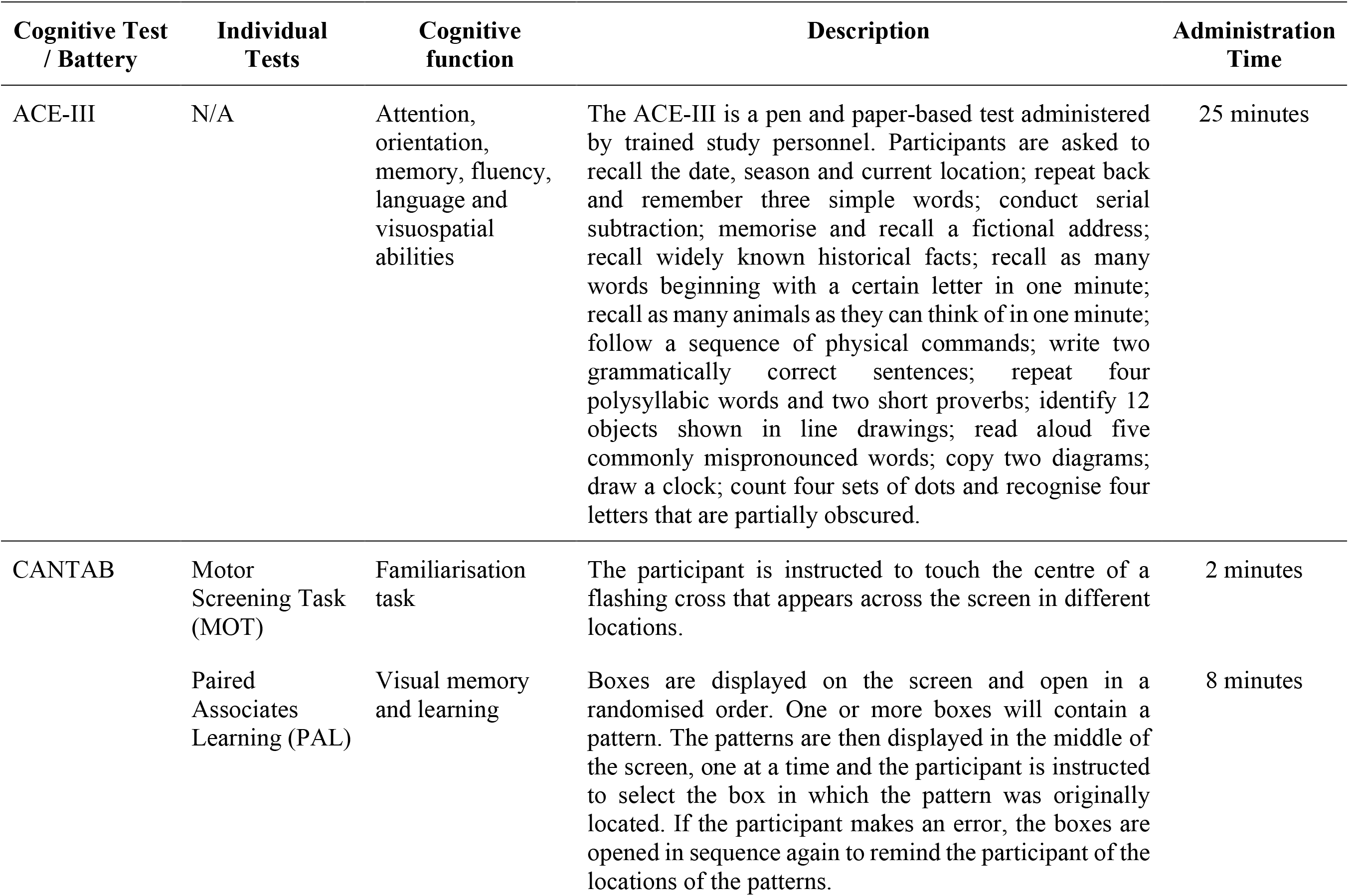
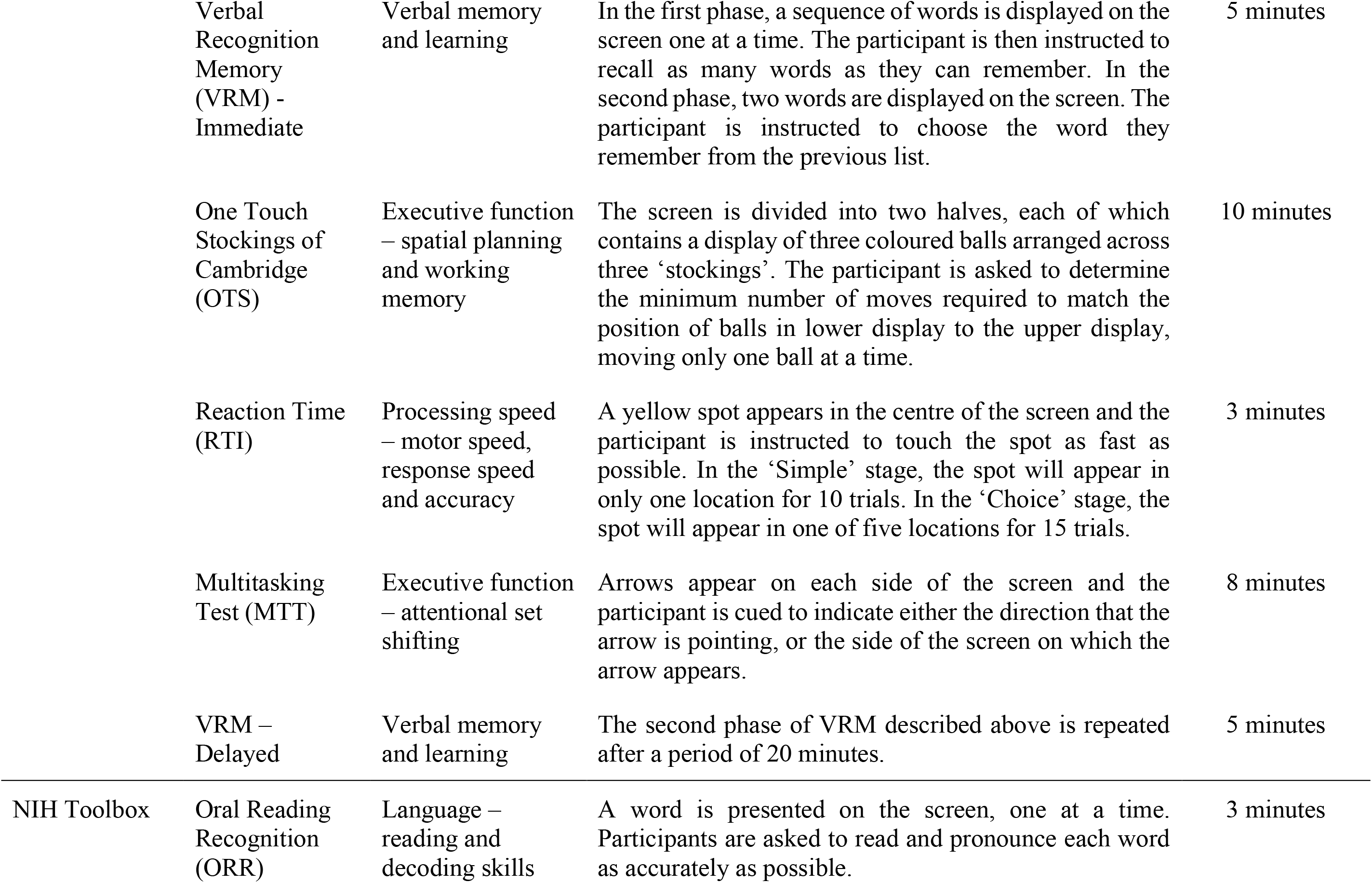
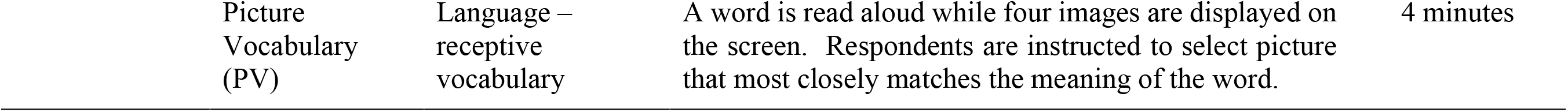
Summary of cognitive assessments administered at Baseline, 18 months and 36 months

###### Behavioural and event-related potential (ERP) measures of cognitive control

Participants will complete a cued-trial task-switching paradigm while we record their electroencephalogram (EEG) to assess proactive cognitive control processes [56]. Participants will complete two choice reaction time tasks (letter and number classification) using colour cues that are mapped to each task [57]. The cue validly indicates whether the letter or the number task will be performed on the upcoming target, allowing the participant to upload the task, but not activate a response until after target onset (cue-target interval 1000 ms). EEG will be recorded using a 64-channel system (ANT in Adelaide; BioSemi in Newcastle) while participants complete either each task alone (single-task blocks) or switch between the two tasks using a random cue colour sequence (mixed-task blocks).

Two indices of task-switching performance are derived: switch cost (switch – repeat trials in a mixed-task block) represents the cost of updating task-set on switch trials, whereas mixing cost (repeat trials in a mixed-task vs. a single-task block) represents the additional working memory load of maintaining two or more representations accessible in mixed-task blocks [58, 59]. Increasing age across the adult lifespan is associated with a robust mixing cost, but a less consistent effect on switch cost [60, 61]. Cue-locked ERP analyses show two parietally-maximal difference components: switch-positivity is associated with preparing to shift task-set, whereas mixing-positivity is associated with working memory load. Both positivities vary in amplitude and scalp distribution across the adult lifespan, with age effects on ERPs emerging much earlier than effects on behavioural indices of task-switching [60]. In mid-late life, mixing cost is associated with reduced microstructural integrity of fronto-parietal white matter tracts (ie., superior longitudinal fasciculus, inferior fronto-opercular tract) [62]. Task switching ability has also been shown to be sensitive to variability in physical activity and nutrition [63, 64].

##### Blood pressure

Hypertension is a known modifiable risk factor for dementia [65]. Blood pressure will be assessed using an OMRON Healthcare Co. digital blood pressure monitor (model 1A1B Hem-7000-CIL). Participants will be seated for at least 5 minutes. Three measurements of blood pressure will be taken, each spaced at least one minute apart, with the forearm rested at table height and the cuff will be placed approximately 2–3 cm above the anti-cubital fossa. Both, average values for systolic blood pressure, diastolic blood pressure and heart rate and difference between systolic and diastolic pressure will be calculated.

##### Blood chemistry

Fasting venous blood will be drawn by venepuncture by a trained phlebotomist to measure lipids profiles, glucose, inflammatory markers, erythrocyte fatty acids and genetic risk profiles. A total of 60 mL will be collected, centrifuged (4°C, 4000 rpm, 10 minutes) and frozen at -80°C for batch processing at the end of each data collection timepoint. Blood samples will be analysed in duplicate with commercial assay kits (including quality controls and calibrators) using the KONELAB 20XTi (ThermoFisher, Massachusetts, United States).

##### Alzheimer’s Disease risk

The ANU Alzheimer’s Disease Risk Index (ANU-ADRI) will be administered to calculate risk of Alzheimer’s disease. The ANU-ADRI is an evidence-based self-report measure of exposure to risk and protective factors associated with Alzheimer’s disease (AD), including age, sex, education, body mass index, diabetes, depression, cholesterol, traumatic brain injury, smoking, alcohol intake, social engagement, physical activity, cognitive activity, fish intake, and pesticide exposure. The ANU-ADRI has high predictive validity for incident Alzheimer’s disease and dementia [66].

##### Subjective cognitive decline

Subjective memory complaints and self-reported cognitive change are strong predictors of cognitive decline, even when performance on cognitive tests is unimpaired [67]. Therefore, the Cognitive Function Instrument (CFI) will be administered to capture subjective cognitive decline. The CFI is a 14-item self-report measure of change in daily functioning with reference to functioning one year prior [68]. The CFI has demonstrated high test-retest reliability (ICC: 0.76 – 0.78), and strong correlations with longitudinal Clinical Dementia Rating scores [68].

##### Subjective Sleep Quality

As both objective and subjective measures of sleep are associated with increased risk of cognitive decline and dementia [69], subjective sleep quality will be captured using the Pittsburgh Sleep Quality Index (PSQI). The PSQI is a self-report questionnaire that assesses sleep quality over seven sub scores: ‘Subjective sleep quality’, ‘Sleep latency’, ‘Sleep duration’, ‘Habitual sleep efficiency’, ‘Sleep disturbances’, ‘Use of sleeping medication’, and ‘Daytime dysfunction’. The PSQI has demonstrated high validity when compared with objective measures of sleep quality and disturbance and has high test-retest reliability [70].

##### Quality of life

Quality of life will be assessed using the EuroQol-5 Dimension (EQ5-D). The EQ-5D assesses health-related quality of life across five dimensions: mobility, self-care, usual activities, pain/discomfort and anxiety/depression. The EQ-5D has been validated for use in older populations with cognitive impairment [71]. Further, it is widely used for the economic valuation of health interventions and has Australian-based population norms [72].

#### Covariates

Covariates will include education, socioeconomic status, comorbidities and medications, smoking and alcohol use, body composition, hearing loss and Apolipoprotein E4 (ApoE4).

Education, smoking and alcohol use will be determined from the ANU-ADRI questionnaire [66].

Comorbidities and medication use will be collected through interview questions informed by the Australian Longitudinal Study of Ageing [73]. Participants will be asked to confirm current health conditions, medications and supplements, including medication type, dose, duration of intake, and whether the medication was prescribed by a doctor or health professional.

Body weight will be measured using an electronic scale with volunteers wearing minimal clothing and without shoes. Height will be measured using a wall mounted stadiometer (SECA, Hamburg, Germany) with volunteers in stockinged or bare feet. Waist and hip circumference (in centimetres) will be measured according to the protocols of the International Society for the Advancement of Kinathropometry and used to calculated waist to hip ratio [74].

Hearing loss is a significant predictor of dementia [65] and will be captured using a short questionnaire, previously described Amieva et al [75, 76]. Participants will be asked if they currently wear hearing aids, and if they have experienced any hearing difficulties in the past 12 months. Self-reported hearing loss is highly correlated with audiometric measures [75] and is associated with an increased risk of cognitive decline and dementia over a follow-up period of 25 years [75, 76].

The ApoE4 allele is the strongest genetic risk factor for Alzheimer’s disease [77]. APOE status will be determined through genetic risk profiling of blood samples described above.

### Procedure

Participants will attend assessment visits at Baseline, 18 months and 36 months. Written informed consent will be obtained at the beginning of the Baseline visit at both study sites. All measures will be collected at Baseline and 36 months. Only Session 1 measures will be collected at 18 months. Table 4 lists the measures collected at each session. Due to logistical differences, the order of assessments may vary between Adelaide and Newcastle.

**Table 4.** Schedule of assessments. Session 1, 2 and 3 will be completed at Baseline and 36 months. Only Session 1 will be completed at 18 months.

|  | Measures |
| --- | --- |
| Session 1 | Informed consent<br>Blood pressure assessment<br>Anthropometry<br>Blood sample collection<br>Diet and lifestyle questionnaires (ANU-ADRI, AES, PSQI, EQ5D, CFI)<br>Cognitive assessments (ACE-III, CANTAB, NIH Toolbox)<br>Activity monitor administered |
| Session 2 | MRI<br>Task-switching practice<br>TMS-EEG (Adelaide only)<br>Optical imaging (Newcastle only) |
| Session 3 | Task switching-EEG<br>TMS-EEG (Adelaide only) |

### Analysis

#### Association between changes in 24-h time-use and diet compositions, cognition and brain function

Primary characteristics of the sample will be presented as per STROBE guidelines [78]. Compositional multi-level mixed effects models, testing for interactions between time-point (Baseline, 18 months and 36 months) and compositions of time-use and diet will assess longitudinal associations between dependent (cognition, brain function and structure) and independent (time-use and diet compositions) variables. If significant at p<0.05, interaction terms will be included between the sets of time-use and diet composition log-ratios. Fixed effects will include the time-use and diet compositions (each expressed as a set of isometric log ratios) and covariates. Dietary analyses will also include total energy intake. Random intercepts will account for repeated measures within a person, nested within study sites. A hierarchical nested model structure will allow comparison of linear and non-linear terms for time-point, establishing the model of best fit. Compositional isotemporal substitution will use the above models to quantify estimated change in cognitive function when time is reallocated from one time-use behaviour to another (keeping dietary intake constant), when one food group increases by one serve at the expense of another (keeping time use constant), or when both time-use and diet are changed concurrently.

#### Identifying best days for cognition and brain health

Using iterative application of the compositional isotemporal substitution analyses, we will generate n-dimensional heat maps or response surfaces for within-person change in activity and diet composition and change in outcomes between sets of two time points (consecutive and distal), adjusted for covariates. The highest regions of the response surfaces will correspond to the most beneficial changes for improvement. These “peaks” or contours (e.g., corresponding to best 5% of outcomes) will be expressed in terms of deviations from a reference time-use or diet composition, to provide customised meaningful interpretation in min/d or servings of food groups.

### Power calculation

In the absence of longitudinal data on diet and activity compositions in relation to cognitive outcomes, we powered the ACTIVate study based on cross-sectional pilot data (age: 65 ± 7.6 y, n=83) of a four-part time-use composition associated with ACE-III scores. Multiple regression analyses gave an R^2^ value of 0.20 when including covariates age and sex and compositional variables for activity. We base our power calculation on these results, and assuming three compositional parameters representing a four-part time-use composition (sleep, sedentary time, light physical activity, and moderate-to-vigorous physical activity), and assuming 7 parameters for the covariates (including four compositional parameters for a five-part diet composition). Aiming for 80% power, alpha will be adjusted to 0.01 to account for multiple comparisons. Based on these assumptions, the R package *pwr* [79] estimates a total sample size of 236 is needed for a general linear model. This number is further inflated to account for the longitudinal design, allowing for 25% attrition over the 36-month period, and a 70% response rate at recruitment, resulting in a final sample size of 448 adults at study enrolment (224 per site).

### Data management

All data collected during this study, including informed consent, will be managed using REDCap electronic data capture tools hosted at the University of South Australia. REDCap is a secure, web-based software platform designed to support data capture for research [80, 81]. Where data files are too large to be uploaded to REDCap, files will be stored on a secure online data storage platform hosted at the University of South Australia. Only listed investigators will have access to the data.

### Risks

There are small risks associated with the MRI and TMS non-invasive brain stimulation paradigms for individuals with metal in their head or body and people with a history of epilepsy. To minimise these risks, all participants will be required to complete an MRI and TMS safety screen.

### Ethical considerations

Interested participants will provide written informed consent at study enrolment and verbally re-consent at 18 months and 3 years. If a participant is clinically diagnosed with dementia or develops a disability affecting their ability to consent after Baseline assessments, a study partner will be required and will be asked to provide written informed consent. Study information will still be discussed with the participants and their chosen study partner, and if the participant decides they no longer want to participate further, they will be withdrawn from the study. All participants are free to withdraw from the study at any time with no consequence to them now or in the future.

### Dissemination

The results of the ACTIVate study will contribute to multiple publications and will be presented at several national and international conferences. Data will be published at multiple stages of the study to ensure findings are communicated quickly and directly. In addition, the data for the primary dependent (cognition) and independent variables (diet and activity composition) will be used in the development of a dementia prevention tool.

### Tool creation

Full description of the co-design process involved in digital tool development is beyond the scope of this protocol paper and will be published elsewhere. Briefly, using both the cross-sectional and the longitudinal data collected, we will co-develop a tool with older adults that can be used by older adults and clinicians for dementia prevention. All aspects of the tool interface will be co-designed through an iterative design process. The tool will provide customised suggestions on how to improve time-use and dietary intake. Recommendations will be tailored to an individual’s situation, allowing the user to incorporate non-negotiable constraints on behaviours, for example, long commute times to work. It will also consider constraints due to personal preference, for example, preference to modify diet rather than time-use, or a desire to keep sleep duration constant and modify waking activities only.

### Data sets and availability

Once the ACTIVate study is complete, de-identified data will be made available to other research groups upon reasonable request. To obtain an ethically compliant data set (with de-identified data) researchers will contact the corresponding author and provide evidence of institutional ethics approval for local data storage, analysis and security.

## DISCUSSION

The ACTIVate study is the first of its kind to track time use, diet and cognition in older adults across three successive years with the intention to use emerging statistical analysis techniques (CoDA) to 1) Explore how changes in 24-hour time use and diet are associated with changes in cognition and 2) Identify ‘best days’ for time use and dietary intake that optimise cognition. In addition to the primary aims, each study site is leading an exploratory secondary biological mechanism focus. We tentatively expect that these secondary mechanisms will provide sensitive measures of variation in ‘best day’ compositions over time. A future aim of the ACTIVate study is to use the data collected to co-create with older adults a fully customisable tool that can be rapidly translated into primary care settings and be used for dementia prevention.

### COVID-19 Statement

Data collection for the ACTIVate study was due to begin in March 2020. However, due to the emergence of the novel coronavirus (COVID-19) as a global pandemic and the potential risk posed to older participants, data collection has been postponed until it is deemed safe to continue. Data collection procedures detailed within this paper may be modified to comply with emerging COVID-safe protocols including minimising the time spent in close contact with participants. Further, all investigators will undergo COVID specific training and participants and investigators will wear personal protective equipment as required.

### Conclusions

In the absence of effective pharmacological interventions that prevent dementia, current research efforts are focussed on reducing dementia risk through modifiable lifestyle factors. Indeed, up to 40% of late life dementia is thought to be due to modifiable factors [4]. Further, population estimates suggest that reducing the prevalence of each modifiable dementia risk factor by 10-20% per decade would potentially reduce the worldwide prevalence of Alzheimer’s disease (the most common form of dementia) by between 8.8 and 16.2 million cases [82]. However, to our knowledge, no studies have tracked time use, diet, cognition and brain function (using various experimental neurophysiological techniques) longitudinally across time. Findings from this project are likely to inform new avenues for multidomain interventions that accommodate the co-dependence between time use and diet behaviours for dementia prevention.

## Author Contributions

A.E.S., F.K., M.J.B., K.E.L, T.S.O, M.R.G., M.C.R., D.D., M.F., and J.D. conceptualised the study. A.E.S., A.T.W., M.M., H.A.D.K. and F.K. developed the cognitive measures. A.E.S., D.D. and T.S.O. developed the activity measures. A.T.W., C.E.C. and K.J.M. developed the dietary measures. M.J.B. and B.P. developed the MRI measures. A.E.S., M.R.G. and M.L.M. developed the TMS measures. A.E.S., M.R.G., M.L.M., M.M., M.A., F.A. and F.K. developed the EEG measures. M.F., G.G., M.A., F.A. and F.K. developed the optical imaging measures. T.S.O, D.D. and J.D. developed the statistical approach. A.E.S., K.E.L., D.D. and R.T.S will oversee the development of the translation tool. A.E.S and A.T.W. prepared the manuscript. All authors reviewed manuscript drafts and approved the final version.

## Acknowledgements

We thank Angela Walls and Shiami Luchow for their assistance setting up the Magnetic Resonance Imaging (protocols) at SAHMRI and HMRI, respectively.

## Supplementary Tables and figures

### MRI Supplementary Material

**Supp Table 1.**
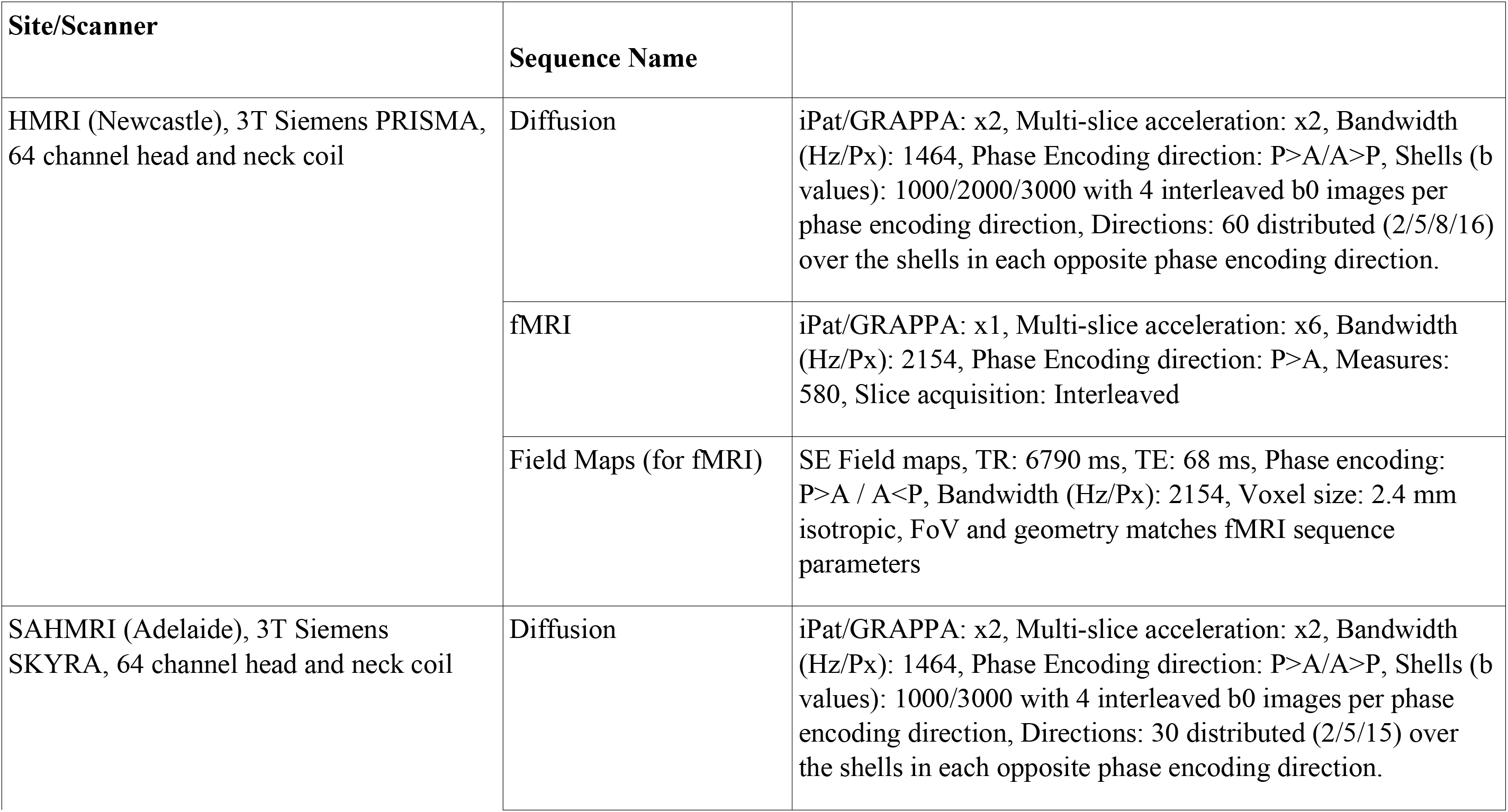
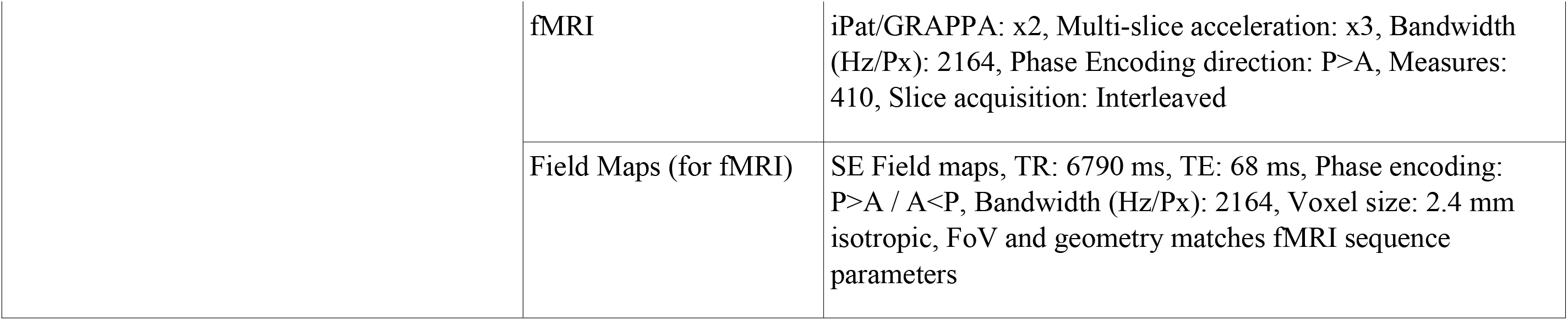
Additional Protocol Information

